# Association between Blood N-3 Fatty Acid Levels and Risk for COVID-19 in the UK biobank

**DOI:** 10.1101/2022.08.19.22278992

**Authors:** William S. Harris, Nathan L. Tintle, Swaminathan Perinkulam Sathyanarayanan, Jason Westra

## Abstract

**Background:** The role of nutritional status and risk for contracting and/or suffering adverse outcomes from SARS-CoV-2 infection is unclear. Preliminary studies suggest that higher n-3 PUFA intakes may be protective.

**Objectives:** The purpose of this study was to compare risk for three COVID-19 outcomes (testing positive for SARS-CoV-2, hospitalization, and death) as a function of baseline plasma DHA levels.

**Methods:** DHA levels (% of total fatty acids) were measured by nuclear magnetic resonance. The three outcomes and relevant covariates were available for 110,584 subjects (hospitalization and death) and for 26,595 ever-tested subjects (positive for SARS-CoV-2) in the UK Biobank prospective cohort study. Outcome data between January 1, 2020 and March 23, 2021 were included. Omega-3 Index (red blood cell EPA+DHA%) values across DHA% quintiles were estimated. Multi-variable Cox-proportional hazards models were constructed and linear (per 1-SD) relations with risk for each outcome were computed as hazard ratios (HRs).

**Results:** In the fully adjusted models, comparing the fifth to the first DHA% quintiles, the HR for testing positive (95% CI) was 0.79 (0.71, 0.89; p<0.001), for being hospitalized was 0.74 (0.58, 0.94; P<0.05), and for dying with COVID-19 was 1.04 (0.69, 1.57; NS). On a per 1-SD increase in DHA% basis, the HRs were: for testing positive, 0.92 (0.89,0.96; p<0.001); for hospitalization, 0.89 (0.83, 0.97; p<0.01); and for death, 0.95 (0.83,1.09). Estimated Omega-3 Index values across DHA quintiles ranged from 3.5% (quintile 1) to 8% (quintile 5).

**Conclusions:** These findings suggest that nutritional strategies to increase circulating n-3 PUFA levels, such as increased consumption of oily fish and/or use of n-3 fatty acid supplements, may reduce risk for adverse COVID-19 outcomes.

## Introduction

The COVID-19 pandemic has arguably been the most disruptive worldwide event since World War II. Its impact was felt not only in the health sector (morbidity and mortality), but also in the economic, social and political realms. A tremendous amount of research has been conducted on the causes of and treatments for the infection, including efforts to discover the factors that affect susceptibility to the disease. In addition to demographic and physiologic characteristics (e.g., obesity, age, underlying health conditions, socio-economic status, etc.), nutritional considerations have been explored. Nutrients such as zinc, vitamins C and D, as well as dietary bioactives (e.g., probiotics) have been suggested as possible protective agents against SARS-CoV-2 infection and/or COVID-19 sequala(1). In addition to these, the long-chain n-3 fatty acids of marine origin – EPA and DHA - have also been proposed to be protective against COVID-19(2-5). DHA and EPA are the main precursors for inflammation resolving mediators, including maresins, resolvins and protectins (6). These mediators downregulate the production of cytokines and also improve macrophage-mediated removal of inflammatory debris, microbes and promote apoptosis of neutrophils(2). Accordingly, higher levels of EPA and DHA in the tissues could reduce the severity of the inflammatory response to SARS-CoV-2 infection. Small (<100 patients) studies have reported that higher red blood cell levels of EPA+DHA (i.e., the Omega-3 Index, O3I(7)) at admission were associated with lower risk for adverse COVID-19 outcomes(8-10), and pilot intervention studies have reported possible benefits from n-3 supplementation(11-13). The availability of plasma DHA levels in a subset of individuals in the UK Biobank cohort(14) affords the opportunity to explore the relationships between n-3 biostatus and risk for COVID-19 outcomes. Two prior studies on this topic have been published. Julkunen et al.(15) and Sun et al.(16) both recently reported that higher DHA levels, among several other biomarkers, were associated with lower risk for infection with or hospitalization with COVID-19. In addition, reported use of fish oil supplements was recently linked with lower risk for COVID-19 in the UK cohort(17). The purpose of the present study was to expand upon the work of these investigators by 1) examining the n-3 relations with additional COVID-19 outcomes (i.e., death), 2) performing more granular analyses (i.e., quintiles), 3) exploring the effects of several covariates on these relationships, and 4) translating n-3 biostatus into the O3I, a more commonly used measure of n-3 status.

## Methods

### Participants

UK Biobank is a prospective, population-based cohort of approximately 500,000 individuals recruited between 2007 and 2010 at assessment centers across England, Wales and Scotland. Baseline data derived from questionnaires, biological samples and physical measurements were collected on all participating individuals, with longitudinal monitoring occurring via a mix of in-person and Electronic Medical Record data(14, 18). The participants completed a touchscreen questionnaire, which collected information on socio-demographic characteristics, diet, and lifestyle factors. Anthropometric measurements were taken using standardized procedures. The touchscreen questionnaire and other resources are shown on the UK Biobank website (http://www.ukbiobank.ac.uk/resources/). UK Biobank has ethical approval (Ref. 11/NW/0382) from the North West Multi-centre Research Ethics Committee as a Research Tissue Bank. This approval means that researchers do not require separate ethical clearance and can operate under the Tissue Bank approval. All participants gave electronic signed informed consent. The UK Biobank study was conducted according to the guidelines laid down in the Declaration of Helsinki. The UK Biobank protocol is available online (http://www.ukbiobank.ac.uk/wp-content/uploads/2011/11/UK-Biobank-Protocol.pdf).

A total of 117,946 individuals from the UKbiobank cohort had blood fatty acid data (**Supplementary Flow Diagram**). We first, dropped people who died before January 1, 2020 (i.e., before the pandemic began), leaving a sample of N=111,240 individuals. After dropping a small number of individuals with missing covariates (listed below under Statistical methods) our final analysis dataset consisted of N=110,584.

### Exposure

The primary exposure was plasma DHA (expressed as a percent of total plasma fatty acids) obtained from blood samples collected at time of enrollment using by nuclear magnetic resonance (Nightingale Health Plc, Helsinki, Finland(19)). In order to estimate the O3I associated with these DHA levels, we used an equation described by Schuchardt et al.,(20) which found a correlation between the observed and estimated O3I (eO3I) of 0.83.

### Outcomes

We examined three primary endpoints: (1) death from COVID, (2) hospitalization with confirmed SARS-CoV-2 infection, and (3) having tested positive for SARS-CoV-2. Data were available on these three outcomes through March 23, 2021 including the outcome itself (yes/no) and the date for which the outcome was recorded. Any individuals recorded as having died and/or been hospitalized with COVID were imputed as having tested positive for COVID if not already noted as such in the dataset (N=174, 4% of the total). Because of inconsistencies in the reporting of testing data across regions of the UK (e.g., testing data were not available for Wales), and to provide a more precise outcome measure, the analysis sample for the ‘tested positive’ outcome, consisted of only individuals for whom a test result was available (e.g., ever tested; N=26,597). Links to the UK Biobank outcomes, exposures and covariates used here are given in **Supplementary Table 6**.

### Statistical Methods

Cox-proportional hazards models were used to predict all three outcomes. For each outcome, four separate models were run predicting the outcome using DHA% while using different sets of covariates with prior evidence of association with confirmed infection with SARS-CoV-2(21). Model 1 unadjusted; Model 2 adjusted for age at start of pandemic, sex, and race (self-identified); Model 3 included Model 2 covariates + waist circumference; and Model 4 included Model 3 covariates + Townsend Deprivation Index (a measure of relative socio-economic status), time since enrollment, smoking status, education, self-reported health, blood pressure medications, slow walking pace, fresh fruit, dried fruit, salad/raw vegetables, cooked vegetables and grain fiber (from cereals and breads (22)). For models with sets of covariates which substantially attenuated hazard ratios (particularly Models 2 vs 3), the relative contribution of each individual variable to the explained variation was evaluated by a drop-one analyses. The effect of dropping each individual covariate illuminates the relative impact of each on the model concordances. Other covariates identified in prior studies(21) for test positivity included cystatin-C, HbA1c, forced expiratory volume, and HDL cholesterol. However, these variables were missing for ≈17% of subjects, thus, we only included them in a sensitivity analysis (Model 5 = Model 4 covariates + these four variables). As in prior studies, we focused primarily on modeling the relationships between DHA% and COVID outcomes with quintiles and continuous linear relationships. In secondary analyses we further investigated potential non-linearities using cubic splines. Cubic splines were compared to linear models using nested ANOVA to test for significant model improvement. All analyses were conducted in R version 4.1.2. and used a 0.05 statistical significance threshold.

## Results

Descriptions of the analytic cohorts are presented in **Table 1**. Among the 110,584 people included in this study, the average age at the beginning of the pandemic was about 68 yrs, the vast majority were White and a little less than half were males. The average plasma DHA value was 2%, and the eO3I was 5.6%. Less than 1% of the full cohort were hospitalized with COVID, and about 20% of those hospitalized died from it. Among the 26,597 people in the dataset who ever got tested for SARS-CoV-2 infection, about 15% tested positive at some point between January 1, 2020 and March 23, 2021. In those who got tested, plasma DHA% was similar to that of the whole cohort. When divided into quintiles, the plasma DHA% ranged from <1.49% in quintile 1 to >2.50% in quintile 5 (**Tables 2-4**). The median eO3I values corresponding to each plasma DHA% quintile ranged from 3.54% to 7.96% in quintiles 1 and 5, respectively (**Figure 1**).

**Table 1.** Distribution of analysis variables^1^.

|  | **All** | **Ever tested for COVID** |
| --- | --- | --- |
| N | 110,584 | 26,597 |
| Sex - Male | 49,638 (44.9%) | 12,368 (46.5%) |
| Race - White | 104,340 (94.4%) | 25,038 (94.1%) |
| Positive tests<sup>2</sup> | 4081 (3.7%) | 4081 (15.3%) |
| Hospitalized with COVID<sup>2</sup> | 837 (0.8%) | 837 (3.1%) |
| Dead with COVID<sup>2</sup> | 235 (0.2%) | 235 (0.9%) |
| Age (on January 1, 2020) | 68.1 (8.1) | 68.6 (8.2) |
| Years since enrollment | 11.39 (0.88) | 11.39 (0.89) |
| Estimated<sup>3</sup> Omega-3 Index (%) | 5.6 (1.89) | 5.56 (1.91) |
| Plasma DHA (% of total fatty acids) | 2.02 (0.68) | 2 (0.68) |
| Waist Circumference (cm) | 90 (13.29) | 91.23 (13.74) |
| BMI (kg/m<sup>2</sup>) | 27.38 (4.73) | 27.77 (4.93) |
| Townsend Deprivation Index<sup>4</sup> | -1.35 (3.07) | -1.17 (3.19) |
| Education - College/University | 42,106 (38.1%) | 9,657 (36.3%) |
| Smoking status - Never | 61,236 (55.4%) | 13,822 (52%) |
| Self-report health - Excellent | 18,606 (16.8%) | 3,946 (14.8%) |
| Long standing illness - None | 74,205 (67.1%) | 16,325 (61.4%) |
| Blood pressure medications - Yes | 4844 (4.4%) | 1398 (5.3%) |
| Diabetes - Yes | 5339 (4.8%) | 1629 (6.1%) |
| Walking pace – Slow<sup>5</sup> | 7943 (7.2%) | 2483 (9.3%) |
| Fresh fruit (pieces per day) | 2.23 (1.58) | 2.23 (1.65) |
| Dried fruit (pieces per day) | 1.89 (2.29) | 1.88 (2.37) |
| Fresh Vegetables/Salad (heaped tbsp per day) | 2.16 (2.17) | 2.14 (2.15) |
| Cooked Vegetables (heaped tbsp per day) | 1.9 (2.51) | 1.91 (2.52) |
| Grain fiber (estimated g/week) | 21.76 (14.52) | 21.34 (14.32) |

1. N (percent) or mean (SD)
2. Between January 1, 2020 and March 23, 2021
3. Based on Schuchardt et al.(20)
4. An index of 0 means 'average deprivation' in the UK; a negative value means less deprived than average and a positive means more deprived than average.
5. <3 miles per hr ("steady" is 3-4 and "fast" is >4)

**Table 2.** Hazard ratios (HRs, 95% CI) from Cox-proportional hazards models by quintile for testing positive for SARS-CoV-2 as a function of plasma DHA% among those who were ever tested (n=26,597)

| Plasma DHA% Quintile (Q; min, max) | Unadjusted | Model 2<sup>1</sup> | Model 3<sup>1</sup> | Model 4<sup>1</sup> |
| --- | --- | --- | --- | --- |
| Q1 (<1.49; reference) | 1 | 1 | 1 | 1 |
| Q2 (1.49, 1.79) | 0.94 (0.86,1.03) | 0.96 (0.88,1.05) | 0.99 (0.9,1.08) | 0.99 (0.91,1.09) |
| Q3 (1.80, 2.09) | 0.86 (0.78,0.94)** | 0.90 (0.82,0.99)* | 0.95 (0.86,1.04) | 0.97 (0.89,1.07) |
| Q4 (2.10, 2.50) | 0.73 (0.66,0.8)*** | 0.79 (0.72,0.87)*** | 0.85 (0.76,0.93)** | 0.89 (0.80,0.98)* |
| Q5 (>2.50) | 0.60 (0.55,0.67)*** | 0.68 (0.61,0.76)*** | 0.74 (0.67,0.82)*** | 0.79 (0.71,0.89)*** |
| Linear across quintiles | 0.89 (0.87,0.9)*** | 0.91 (0.89,0.93)*** | 0.94 (0.91,0.96)*** | 0.95 (0.92,0.97)*** |
| Linear trend per SD | 0.84 (0.81,0.86)*** | 0.87 (0.84,0.9)*** | 0.90 (0.87,0.93)*** | 0.92 (0.89,0.96)*** |

<sup>1</sup>Model 2: adjusted for age at start of pandemic, sex, and race; Model 3: Model 2 covariates + waist circumference; Model 4: Model 3 covariates + Townsend Deprivation Index, time since enrollment, smoking status, education, Self-reported health, blood pressure medications, slow walking pace, fresh fruit, dried fruit, fresh vegetables, cooked vegetables and grain fiber. \* p<0.05; \*\* p<0.01; \*\*\* p<0.001.

**Figure 1.**
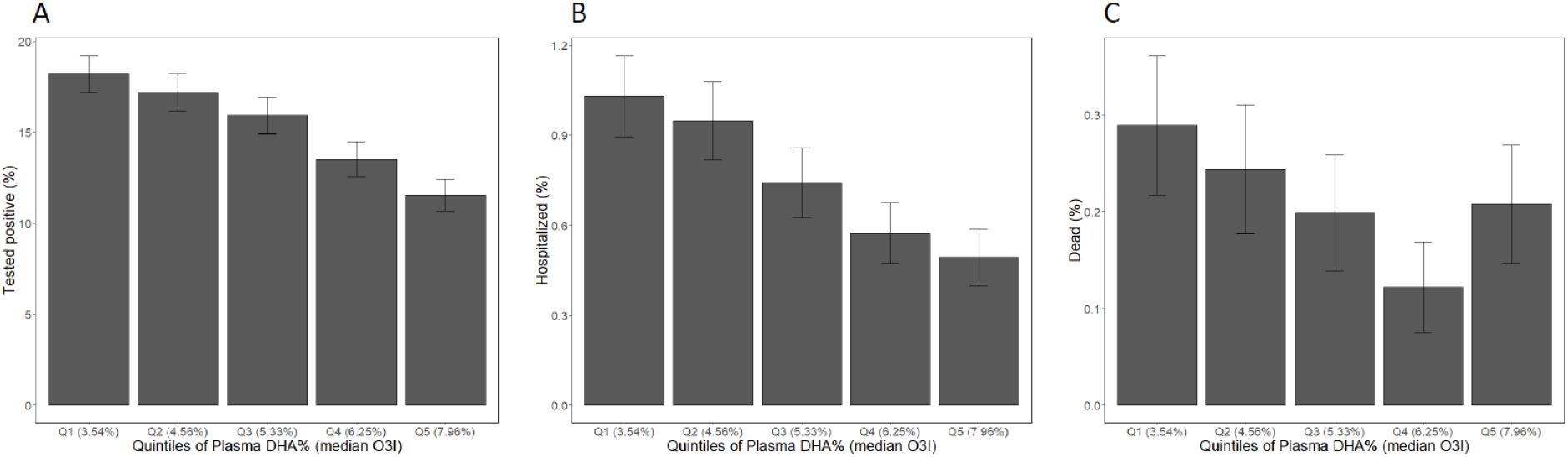
The percent of individuals experiencing COVID-19 outcomes between January 1, 2020 and March 23, 2021 by quintile of the plasma DHA%. The median estimated Omega-3 Index of each quintile is also shown. A, testing positive for SARS-CoV-2 (n=26,597); B, being hospitalized with COVID-19 (n=110,584); and C, dying with COVID-19 (n=110,584).

There was a strong, inverse and dose-related relationship between plasma DHA% and risk for having a positive SARS-CoV-2 test (**Figure 1A**) which was partially attenuated with each level of multivariable adjustment (**Table 2**). Nevertheless, in the fully adjusted model, individuals in quintile 5 were 21% less likely to test positive than those in quintile 1 (p<0.001), and the risk for a positive test was 8% lower for each 1-SD increase in plasma DHA% (p<0.001).

As regards hospitalization with COVID, again there was again a significant, inverse association with the plasma DHA % which was partially attenuated with increasing adjustment (**Figure 1B**). In the fully adjusted model (**Table 3**), individuals in quintile 5 were at 26% lower risk for hospitalization than those in quintile 1 (P<0.05), and risk was 11% lower per 1-SD higher in DHA% (p<0.001).

**Table 3.** Hazard ratios (HRs, 95% CI) from Cox-proportional hazards models for hospitalization with COVID-19 as a function of plasma DHA% (n=110,584)

| Plasma DHA% Quintile (Q; min, max) | Unadjusted | Model 2<sup>1</sup> | Model 3<sup>1</sup> | Model 4<sup>1</sup> |
| --- | --- | --- | --- | --- |
| Q1 (<1.49; reference) | 1 | 1 | 1 | 1 |
| Q2 (1.49, 1.79) | 0.92 (0.76,1.11) | 0.95 (0.79,1.15) | 1.07 (0.88,1.29) | 1.11 (0.92,1.34) |
| Q3 (1.80, 2.09) | 0.72 (0.59,0.88)** | 0.74 (0.61,0.91)** | 0.90 (0.74,1.11) | 0.97 (0.79,1.19) |
| Q4 (2.10, 2.50) | 0.56 (0.45,0.69)*** | 0.56 (0.45,0.7)*** | 0.73 (0.59,0.92)** | 0.81 (0.65,1.02) |
| Q5 (>2.50) | 0.48 (0.38,0.6)*** | 0.46 (0.36,0.58)*** | 0.63 (0.5,0.8)*** | 0.74 (0.58,0.94)* |
| linear-trend-quintiles | 0.82 (0.78,0.86)*** | 0.82 (0.78,0.86)*** | 0.89 (0.84,0.93)*** | 0.92 (0.87,0.97)** |
| linear-trend-per SD | 0.75 (0.7,0.81)*** | 0.75 (0.69,0.81)*** | 0.84 (0.78,0.91)*** | 0.89 (0.83,0.97)** |

Model 2: adjusted for age at start of pandemic, sex, and race; Model 3: Model 2 covariates + waist circumference; Model 4: Model 3 covariates + Townsend Deprivation Index, time since enrollment, smoking status, education, Self-reported health, blood pressure, slow walking pace, fresh fruit, dried fruit, fresh vegetables, cooked vegetables and grain fiber. \* p<0.05; \*\* p<0.01; \*\*\* p<0.001

Finally, for death from COVID, relations with plasma DHA% were more complex (**Figure 1C, Table 4**). In all models, risk for death was significantly lower in quintile 4 vs quintile 1 (p<0.05), but in quintile 5, the risk reduction was partially attenuated and not significant in three of the four models (the exception being model 2 where risk was 39% lower, p<0.05). In the unadjusted model and model 2, risk for death was 17% and 22%, respectively, lower (p<0.05) per 1-SD increase in DHA%. However, in the fully adjusted model, it was attenuated to non-significance (Table 4). Also in this model, the difference between risk for death in quintiles 4 and 5 was significant (p=0.04).

**Table 4.**
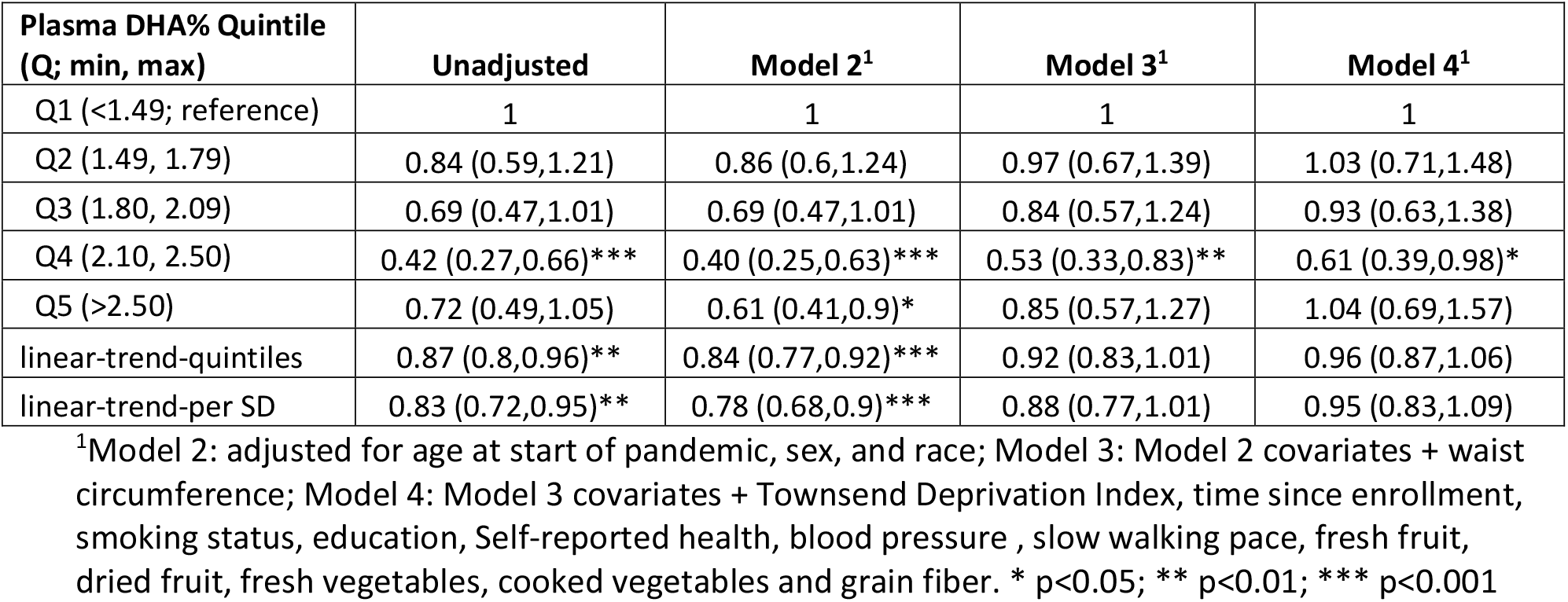
Hazard ratios (HRs, 95% CI) from Cox-proportional hazards models for death with COVID-19 as a function of plasma DHA% (n=110,584)

As mentioned for all three outcomes, there was some attenuation of the relationship between DHA% and risk with adjustment for covariates. Most of the attenuation in risk, particular for hospitalization and death, was seen in model 3, which added waist circumference as a covariate in addition to age, sex and race [from model 2)]. It was therefore of interest to examine each of these four covariates to see the extent to which each was responsible the observed attenuation in relationship. This was done by computing the concordance between DHA% and each of the three outcomes, and then systematically removing each of the variables from the model and calculating the resultant drop in concordance. The results of this exercise are shown in **Table 5**. Regardless of the outcome, the removal of sex and then race from the covariate list had little effect on the concordance. Age and waist circumference were the more important covariates. For testing positive, the inclusion of age had the greatest impact on the observed association with DHA%, decreasing the concordance by 45%. Comparatively, the removal of waist circumference reduced the concordance by only 3%. For hospitalization, waist circumference was the more influential, reducing concordance by 19% compared to 10% for age. For death, age was again the major factor followed by waist circumference, reducing the concordances by 32% and 8%, respectively. Finally, there was some additional attenuation in model 4 (vs model 3) when 12 more covariates were included in the analyses (**Table 4**).

**Table 5.** Effects of the removal of age, sex, race and waist circumference from model 3 on model concordance for the COVID-19 outcomes of interest.

|  | Concordance | Absolute Decrease |  |  |  | Percent Decrease* |  |  |  |
| --- | --- | --- | --- | --- | --- | --- | --- | --- | --- |
|  |  | Age | Sex | Waist circumference | Race | Age | Sex | Waist circumference | Race |
| Positive test | 0.6298 | 0.0584 | 0.0001 | 0.0038 | 0.0042 | 45.0% | 0.0% | 3.0% | 3.0% |
| Hospitalization | 0.7047 | 0.0213 | 0.0011 | 0.0379 | 0.0121 | 10.0% | 1.0% | 19.0% | 6.0% |
| Death | 0.8088 | 0.0998 | 0.0011 | 0.024 | 0.0181 | 32.0% | 0.0% | 8.0% | 6.0% |

\* Calculated by dividing the absolute decrease by the concordance - 0.5

As a sensitivity analysis we included a fifth model with diminished sample sizes (owing to missing covariate data, **Supplementary Table 1**) that additionally adjusted for cystatin-C, HbA1c, forced expiratory volume, and HDL cholesterol. The inclusion of these variables (and the loss of 16.5% of the sample owing to missing data) resulted in further attenuation of the DHA% vs. outcome relationships (**Supplementary Tables 2-4**). Finally, secondary analyses using spline models did not significantly improve upon the continuous models for any outcome and for any level of adjustment (data not shown).

As noted, there were monotonic inverse relations between DHA% and risk for testing positive and for hospitalization. This was also true for death through the fourth quintile, but the trend reversed for the highest quintile. Thus, it was of interest to explore the differences in risk factor and demographic profiles across the quintiles of DHA% among the 235 individuals who died to see if some characteristic of those in quintile 5 might help explain this reversal, especially vs quintile 4 (where the favorable trend was still evident, **Supplementary Table 5**). The most obvious (and only statistically significant) difference between quintiles 5 and 4 was in reported fish oil supplement use, with 22% of the former reporting use and 61% of the latter (p=0.002). This naturally explains why this group had the highest DHA levels. Quintile 5 subjects were also less likely to be white than quintile 4 subjects (74% vs 93%, p=0.1) and exercised less, but also tended to be younger and less obese.

## Discussion

The purpose of this study was to examine the relationships between plasma DHA% at baseline in the UKBB and three COVID-19 related outcomes. We found that a 1-SD higher DHA% was associated with an 8% lower risk for testing positive for SARS-CoV-2 and an 11% lower risk for hospitalization in the multivariable adjusted models. The relationship with risk for death with COVID-19 was also lower (5% lower per 1-SD of DHA%), but it was not statistically significant. Thus, a higher n-3 level appeared to be protective against both getting infected with the virus and with having a severe enough infection to require hospitalization. Regarding risk for death with COVID-19, across the first four DHA% quintiles, risk decreased in a strongly dose-related manner. However, in the highest DHA quintile, the trend reversed, and the difference (from quintile 1) was no longer significant. Beyond the quintile 5 enrichment in subjects reporting fish oil supplement use and in non-white participants, the reasons for this partial reversal are unclear; but caution should be used given the small number of deaths (n=235). Finally, we were able translate DHA% quintiles into the more familiar Omega-3 Index, and we found that the highest risk (i.e., quintile 1) eO3I value was around 3.5%, whereas the lowest risk (quintile 5) value was around 8%. These values comport well with the O3I risk cut points [originally proposed in 2004 for death from cardiovascular disease (7)] of <4% (high risk) and >8% (low risk) and imply that these target levels apply to COVID-19 outcomes as well.

In general, our findings support the previous work of Julkunen et al. who reported on the relations between risk for severe (i.e., hospitalization with) COVID-19 and multiple circulating biomarkers in the UK Biobank(15). They found that a 1 SD increase in DHA% was associated with about a 23% reduction in risk for hospitalization in an age- and sex-adjusted model (compared to 25% here in Model 2). They did not adjust for any other covariates, however, nor did they report on either test positivity or mortality. Sun et al.(16) also published on the relationships between multiple fatty acid metrics and COVID-19 outcomes in the UKBB. Our report also differs from theirs in several ways. First, we were able to translate NMR plasma FA data to the eO3I, a metric that has been widely used clinically to risk stratify patients for a variety of conditions(23-29). Second, although Sun et al. adjusted for more variables that Julkunen et al. did, their list included only age, sex, ethnicity, BMI, Townsend deprivation index, and assessment center. As noted, we adjusted for additional variables including those reported by Ho et al.(21) to be significant determinants of risk for testing positive in hospital for SARS-CoV-2 and those reflecting better dietary habits. Third, like Julkunen et al., Sun et al. did not examine risk for death. Finally, uniquely compared to others, we have reported quintile level relationships and explored non-linear relationships between DHA% and COVID-19 outcomes. The latter could help define optimal or target blood O3I levels potentially useful in clinical practice.

Other reports from within the UKBB have described significant inverse relationships between reported fish oil supplement use [which is associated with a higher O3I] and risk for death from all causes, risk for incident cardiovascular events(30) as well as primary liver cancer(31), inflammatory bowel disease(32), dementia(33), and as noted earlier, COVID-19(17).

Outside of the UKBB, the associations between the O3I or other blood n-3 biomarkers and a variety of COVID-19 related outcomes in small studies (<100 patients each) were reported. We(8) and a research group in Chile(9, 10) have both shown that higher O3I levels were associated with better clinical outcomes. A study in 26 children with multi-system inflammatory syndrome linked to COVID-19 infection found lower levels of EPA and DHA compared with historical controls(34). Two double-blind, randomized, controlled trials (RCTs) used n-3 PUFAs to treat COVID-19. Doaei et al. compared an EPA+DHA enriched enteral nutrition product (n=28) with regular enteral nutrition (n=73) critically ill patients hospitalized with COVID-19(13) and reported a higher 1-month survival rate, better renal function, improved lymphocyte counts and better arterial blood gas parameters. Sedhigiyan et al. compared hydroxychloroquine treatment to hydroxychloroquine plus n-3 FAs and found that patients randomized to the latter regimen had less myalgias and fatigue, and lower erythrocyte sedimentation rates and CRP levels(35).

### Mechanisms

There are several mechanisms that could, in theory, explain the relationships observed here. Beginning with the reduced risk for positive infection, the SARS-COV-2 virus is known to infect the host cells via binding of the spike (S) glycoprotein with the surface ACE2 receptors, expressed in the pneumocytes and other cell types. An *in silico* study by Vivar-Sierra et al. found that DHA and EPA also had the potential to hold the S glycoprotein in a closed confirmation(36). These findings were confirmed by Goc et al.(37) who studied the effect of EPA on host cellular proteases such as transmembrane protease serine protease-2 (TMPRSS-2) and cathepsins that cleave spike proteins and enable cellular entry. EPA effectively inhibited the enzymatic activity of cathepsin L and TMPRSS2. Finally, n-3 PUFAs may also play a role in intracellular mechanisms of inhibiting viral replication. MERS-CoV is known to reprogram the Sterol Regulatory Element Binding Protein (SREBP) dependent lipogenesis pathway to ensure its replication(38). N-3 fatty acids are intracellular inhibitors of SREBP transcription and maturation and may play role in counteracting the viral activation of SREBP1/2 thus decreasing viral replication. N-3 fatty acids are known to have anti-inflammatory actions via alterations in membrane physio-chemistry which alter the activity of toll-like receptor 4, diminishing signal transduction to nuclear factor kappa B which in turn blunts the signaling cascade that produces inflammatory cytokines and adhesion molecules(39). In addition, n-3 fatty acids serve as substrates for the production of a wide array of oxylipins (prostaglandins, leukotrienes, and inflammation resolving mediators) that work in harmony not only suppress inflammatory pathways, but also actively promote the resolution of inflammation (for reviews see (2-4, 40, 41)). Taken together, there is a strong scientific rationale for a favorable effect of higher tissue n-3 levels and protection against both infection with SARS-CoV-2 and its downstream sequelae.

### Strengths and weakness

The greatest strength of this study was the ability to query the UKBB data base with >110,000 individuals with baseline n-3 FA levels measured and then followed for >10 years. Second, we were able to link blood n-3 levels with three important COVID-19 related outcomes: positive testing, hospitalization and death. We also considered several more relevant covariates in our modeling than past authors have included reducing the chances for – but by no means eliminating - unmeasured confounding. HRs for each outcome were not materially affected by the inclusion of fruit, vegetable and fiber intakes as covariates. This suggests that higher DHA levels are not simply a surrogate marker of a healthier diet. Finally, we translated nuclear magnetic resonance -derived n-3 metrics into the more familiar and clinically useful O3I metric.

As to limitations, the first is one common to of all studies linking circulating fatty acid levels measured at baseline with disease outcomes occurring many years later: how stable are blood n-3 FA levels over time? How confident can we be that levels measured at baseline were the same at the outbreak of the pandemic. In the Framingham Offspring cohort, we found that within-persons, the RBC-based O3I was stable between 1999 and 2006 in subjects who did not initiate fish oil supplementation; naturally it increased in those who did(42). Other studies have examined plasma fatty acid stability over time and reported pre-to-post correlations of between 0.5 and 0.8(43-46). Importantly, when baseline levels and follow-up levels were each used to predict risk for incident heart failure in one study(46), both metrics were equally predictive. In the UKBB cohort, baseline (2006-2010) fish oil supplement use was reported by 31% and the consumption of at least 2 meals per week of oily fish by 18%(20). At a random repeat visit between 2012-2013 in 20,334 subjects, supplement use was reported by 32.7%, and >1 oily fish serving per week by 20%. Hence it is reasonable to assume that since the two major determinants of blood n-3 levels remained stable, that baseline n-3 biomarker levels generally reflected levels later in life. Indeed, any major shifts in DHA% after baseline would likely have biased our results towards the null, suggesting that the relationships detected may in fact be stronger than they appear. Another potential limitation is the impact of vaccination status on our findings. The study period covered by this report was from January 1, 2020 (herein defined as the beginning of the pandemic) to March 23, 2021. By the latter date, about 38% of the UK population had received at least one dose of the vaccine, but only 2.6% had received two doses(47). Vaccination has been shown to not only reduce the severity of COVID-19 once contracted, but also to lower the risk for testing positive(48).

Unfortunately, data on vaccination status were not available for this analysis, but in order for differential vaccination status to have biased our results, one would have to posit that people with higher n-3 levels were more likely to have been vaccinated than those with lower levels during the approximately 3-4 month period of this study for which vaccines were available (Dec 2020-March 2021). Although this is plausible [i.e., higher n-3 levels are associated with higher SES and healthful behaviors and exercise and a greater consumption of oily fish and fish oil supplements], we are unable to confirm or deny this hypothesis.

Finally, by its very nature, the UKBB is not a random selection of all individuals in the UK; it is limited to those in a fixed age range who agreed to participate. Thus, the results should not be generalized beyond that context, and outcome data (all taken from electronic medical records) were not formally adjudicated, potentially introducing some bias.

## Conclusion

In this study we confirmed the findings of previous investigations that a low n-3 status was associated with increased risk for hospitalization with COVID-19. We extended these findings by also showing reduced risk for testing positive with the virus and by providing evidence that the risk for death may also be reduced. Furthermore, we identified eO3I levels associated with the least (<4%) and greatest (>8%) protection from COVID-19. Taken together, these results suggest that increased intakes of n-3 (from oily fish and/or taking n-3 supplements) should be encouraged to raise the O3I and possibly reduce the risk for COVID-19, and perhaps other, future pulmonary viral infections.

## Data Availability

Codebook and analytic code related to data described in the manuscript will be made available upon reasonable request pending application to and approval by the Fatty Acid Research Institute. Raw data are available via standard application procedures directly from the UKbiobank.

## Abbreviations

CI: confidence interval
eO3I: estimated Omega-3 Index (erythrocyte EPA+DHA%)
HR: hazard ratio

## Acknowledgements

WSH designed the research; NLT conducted the research, analyzed the data and performed statistical analysis; JW assisted in variable and dataset creation; WSH, NLT and SPS wrote the paper; WSH had primary responsibility for the final content. All authors have read and approved the final manuscript.

## Data Sharing

Codebook and analytic code related to data described in the manuscript will be made available upon reasonable request pending application to and approval by the Fatty Acid Research Institute. Raw data are available via standard application procedures directly from the UK Biobank.

