## Supplemental Materials for "Association between Blood N-3 Fatty Acid Levels and Risk for COVID-19 in the UK biobank"

**Flow Chart**

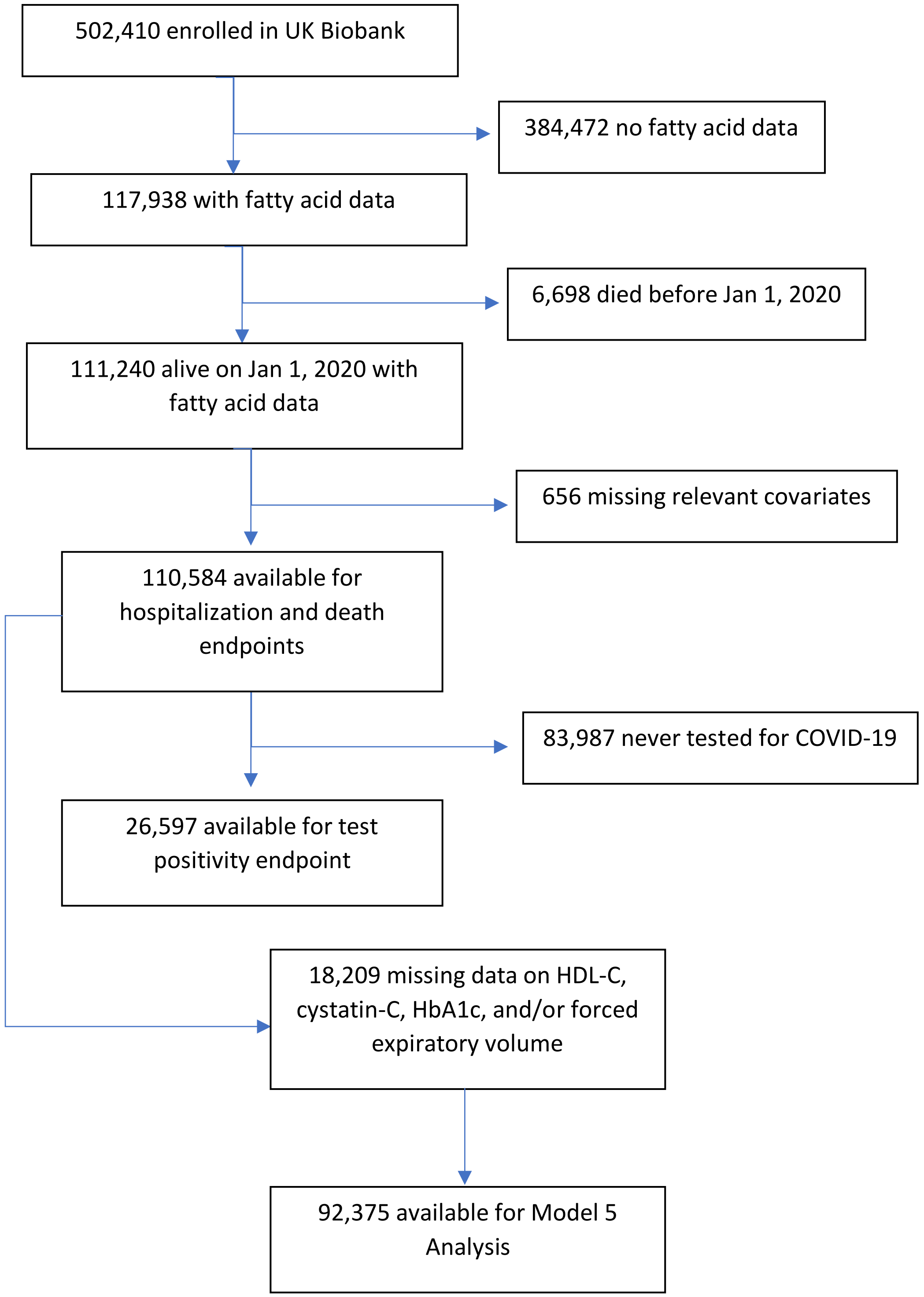

**Supplemental Table 1. Distribution of analysis variables for individuals with data on HDL-C, cystatin-C, HBA1c and forced expiratory volume**

| | **All** | **Ever tested for COVID by 3/23/21** |
|---|---|---|
| N | 92375 | 21951 |
| Sex - Male | 41227 (44.6%) | 10141 (46.2%) |
| Race - White | 87483 (94.7%) | 20739 (94.5%) |
| Positive tests[1] | 3346 (3.6%) | 3346 (15.2%) |
| Hospitalized for COVID[1] | 651 (0.7%) | 651 (3%) |
| Dead from COVID[1] | 187 (0.2%) | 187 (0.9%) |
| Age on January 1, 2020 | 68 (8.1) | 68.4 (8.2) |
| Time since enrollment (yr) | 11.4 (0.87) | 11.4 (0.88) |
| Estimated Omega-3 Index (%) | 5.6 (1.89) | 5.57 (1.9) |
| Plasma DHA (% of total fatty acids) | 2.02 (0.67) | 2 (0.68) |
| Waist Circumference (cm) | 89.73 (13.19) | 90.93 (13.59) |
| Body Mass Index (kg/m$^2$) | 27.3 (4.66) | 27.68 (4.84) |
| Townsend Deprivation Index[2] | -1.4 (3.04) | -1.23 (3.15) |
| Education - College/University | 35437 (38.4%) | 8075 (36.8%) |
| Smoking status - Never | 51596 (55.9%) | 11592 (52.8%) |
| Self-report excellent health | 16158 (17.5%) | 3433 (15.6%) |
| Long standing illness - None | 63274 (68.5%) | 13867 (63.2%) |
| Blood pressure medications - Yes | 3964 (4.3%) | 1133 (5.2%) |
| Diabetes - Yes | 4241 (4.6%) | 1289 (5.9%) |
| Walking pace – Slow[3] | 6046 (6.5%) | 1836 (8.4%) |
| HDL-Cholesterol (mmol/L) | 1.31 (0.32) | 1.3 (0.32) |
| Cystatin C (mg/L) | 0.9 (0.15) | 0.91 (0.17) |
| HbA1c (mmol/mol) | 35.84 (6.36) | 36.21 (6.98) |
| Forced Expiratory Volume (L) | 2.83 (0.79) | 2.8 (0.79) |
| Fresh fruit (pieces per day) | 2.23 (1.58) | 2.24 (1.66) |
| Dried fruit (pieces per day) | 1.89 (2.28) | 1.9 (2.36) |
| Fresh Vegetables/Salad (heaped tbsp per day) | 2.17 (2.17) | 2.15 (2.15) |
| Cooked Vegetables (heaped tbsp per day) | 1.9 (2.51) | 1.93 (2.5) |
| Grain fiber (est g/week) | 21.79 (14.46) | 21.39 (14.2) |

**Supplemental Table 2. Testing positive for COVID-19 among those who ever tested in five models (n=21,951)**

| Prevalence | Unadjusted | Model 2 | Model 3 | Model 4 | Model 5 |
|---|---|---|---|---|---|
| 18.0% (832/4612) | 1 | 1 | 1 | 1 | 1 |
| 16.4% (723/4416) | 0.9 (0.81,1)* | 0.92 (0.83,1.02) | 0.95 (0.86,1.05) | 0.96 (0.87,1.06) | 0.97 (0.88,1.08) |
| 16.1% (700/4342) | 0.88 (0.79,0.97)* | 0.93 (0.84,1.03) | 0.98 (0.88,1.09) | 1 (0.91,1.12) | 1.02 (0.92,1.14) |
| 13.4% (565/4224) | 0.73 (0.65,0.81)*** | 0.8 (0.72,0.89)*** | 0.86 (0.77,0.96)** | 0.9 (0.81,1.01) | 0.93 (0.83,1.04) |
| 12.1% (526/4357) | 0.64 (0.58,0.72)*** | 0.73 (0.65,0.82)*** | 0.8 (0.71,0.9)*** | 0.86 (0.77,0.97)* | 0.89 (0.79,1) |
| linear-trend-quintiles | 0.9 (0.88,0.92)*** | 0.93 (0.9,0.95)*** | 0.95 (0.93,0.98)*** | 0.97 (0.94,0.99)* | 0.97 (0.95,1) |
| linear-trend-perSD | 0.85 (0.82,0.88)*** | 0.9 (0.86,0.93)*** | 0.93 (0.89,0.96)*** | 0.95 (0.91,0.99)* | 0.96 (0.92,1)* |
| spline vs. linear | 0.16 | 0.24 | 0.28 | 0.38 | 0.36 |

Model 2 = Age, Sex, Race
Model 3 = Model 2+ Waist
Model 4 = Model 3+ TDI, time since FA, smoking status, education, Self-reported health, BP meds, slow pace, fresh fruit, dried fruit, fresh vegetables, cooked vegetables and grain fiber
Model 5 = Model 4 + Cystatin C, HBA1C, FEV, HDL

**Supplemental Table 3. Hospitalization for COVID-19 in five models (n=92,375)**

| Prevalence | Unadjusted | Model 2 | Model 3 | Model 4 | Model 5 |
|---|---|---|---|---|---|
| 1% (176/18254) | 1 | 1 | 1 | 1 | 1 |
| 0.9% (162/18488) | 0.91 (0.73,1.12) | 0.94 (0.76,1.16) | 1.04 (0.84,1.29) | 1.09 (0.88,1.35) | 1.14 (0.92,1.42) |
| 0.7% (130/18494) | 0.73 (0.58,0.91)** | 0.75 (0.6,0.94)* | 0.9 (0.72,1.14) | 0.98 (0.78,1.24) | 1.05 (0.83,1.32) |
| 0.5% (92/18624) | 0.51 (0.4,0.66)*** | 0.52 (0.4,0.67)*** | 0.67 (0.52,0.87)** | 0.76 (0.58,0.98)* | 0.82 (0.63,1.07) |
| 0.5% (91/18515) | 0.51 (0.39,0.66)*** | 0.49 (0.38,0.63)*** | 0.67 (0.51,0.87)** | 0.79 (0.6,1.03) | 0.88 (0.67,1.16) |
| linear-trend-quintiles | 0.83 (0.78,0.87)*** | 0.82 (0.78,0.87)*** | 0.89 (0.84,0.94)*** | 0.92 (0.87,0.98)** | 0.95 (0.89,1.01) |
| linear-trend-perSD | 0.76 (0.7,0.83)*** | 0.75 (0.69,0.82)*** | 0.85 (0.78,0.92)*** | 0.89 (0.82,0.97)** | 0.93 (0.85,1.01) |
| spline vs. linear | 0.17 | 0.25 | 0.47 | 0.6 | 0.61 |

Model 2 = Age, Sex, Race

Model 3 = Model 2+ Waist

Model 4 = Model 3+ TDI, time since FA, smoking status, education, Self-reported health, BP meds, slow pace, fresh fruit, dried fruit, fresh vegetables, cooked vegetables and grain fiber

Model 5 = Model 4 + Cystatin C, HBA1C, FEV, HDL

**Supplemental Table 4. Death from COVID-19 in five models (n=92,375)**

| Prevalence | Unadjusted | Model 2 | Model 3 | Model 4 | Model 5 |
|---|---|---|---|---|---|
| 0.3% (52/18254) | 1 | 1 | 1 | 1 | 1 |
| 0.2% (39/18488) | 0.74 (0.49,1.12) | 0.76 (0.5,1.16) | 0.85 (0.56,1.29) | 0.9 (0.59,1.37) | 0.93 (0.61,1.41) |
| 0.2% (36/18494) | 0.68 (0.45,1.04) | 0.69 (0.45,1.06) | 0.84 (0.54,1.29) | 0.92 (0.6,1.42) | 0.93 (0.6,1.44) |
| 0.1% (22/18624) | 0.41 (0.25,0.68)*** | 0.4 (0.24,0.66)*** | 0.52 (0.31,0.87)* | 0.61 (0.36,1.01) | 0.62 (0.37,1.05) |
| 0.2% (38/18515) | 0.72 (0.47,1.09) | 0.63 (0.41,0.96)* | 0.87 (0.56,1.35) | 1.04 (0.67,1.63) | 1.09 (0.69,1.73) |
| linear-trend-quintiles | 0.88 (0.8,0.98)* | 0.85 (0.77,0.95)** | 0.93 (0.84,1.03) | 0.97 (0.87,1.08) | 0.98 (0.88,1.09) |
| linear-trend-perSD | 0.85 (0.73,0.99)* | 0.81 (0.69,0.94)** | 0.91 (0.78,1.06) | 0.98 (0.84,1.14) | 0.99 (0.85,1.15) |
| spline vs. linear | 0.33 | 0.44 | 0.59 | 0.67 | 0.69 |

Model 2 = Age, Sex, Race

Model 3 = Model 2+ Waist

Model 4 = Model 3+ TDI, time since FA, smoking status, education, Self-reported health, BP meds, slow pace, fresh fruit, dried fruit, fresh vegetables, cooked vegetables and grain fiber

Model 5 = Model 4 + Cystatin C, HBA1C, FEV, HDL

**Supplemental Table 5. Distribution of demographic and health variables by quintile of plasma DHA% for those who died with COVID-19 (n=235; ranked by p-value comparing Q4 with Q5)**

| | | Q1 | Q2 | Q3 | Q4 | Q5 | Q4 vs. Q5 |
|---|---|---|---|---|---|---|---|
| N | | 64 | 54 | 44 | 27 | 46 | P-value |
| Fish oil supplement use | % | 12.5 | 31.5 | 31.8 | 22.2 | 60.9 | 0.002 |
| White | % | 89.1 | 92.6 | 86.4 | 92.6 | 73.9 | 0.100 |
| HBA1C (mmol/mol) | mean | 44.05 | 40.48 | 40.58 | 37.33 | 40.48 | 0.100 |
| Cystatin C (mg/L) | mean | 1.07 | 1.08 | 1 | 0.93 | 0.97 | 0.240 |
| Exercise > 660 min/wk | % | 18.8 | 18.5 | 20.5 | 37 | 21.7 | 0.250 |
| Age -years | mean | 61.72 | 63.33 | 63.28 | 64.47 | 62.66 | 0.280 |
| Obese | % | 51.60% | 50.00% | 47.70% | 40.70% | 26.10% | 0.300 |
| Male | % | 67.20% | 66.70% | 54.50% | 48.10% | 63.00% | 0.320 |
| Townsend Deprivation Index | mean | 0.2 | 0.2 | 0.9 | -0.65 | 0.07 | 0.400 |
| Stroke before pandemic | % | 7.8 | 7.4 | 9.1 | 3.7 | 10.9 | 0.530 |
| Daily Alcohol use | % | 15.6 | 16.7 | 27.3 | 22.2 | 15.2 | 0.660 |
| Long standing illness | % | 56.3 | 59.3 | 63.6 | 33.3 | 41.3 | 0.670 |
| Urban | % | 93.8 | 92.6 | 100 | 85.2 | 91.3 | 0.670 |
| Rate health poor | % | 6.3 | 11.1 | 20.5 | 11.1 | 6.5 | 0.800 |
| Waist circumference (cm) | mean | 103.75 | 101 | 99.27 | 95.74 | 94.96 | 0.830 |
| Forced expiratory volume (L) | mean | 2.47 | 2.67 | 2.42 | 2.55 | 2.51 | 0.860 |
| Slow walking pace | % | 28.1 | 29.6 | 18.2 | 11.1 | 15.2 | 0.890 |
| Diabetes | % | 18.8 | 20.4 | 25 | 14.8 | 10.9 | 0.900 |
| HDL-C (mmol/L) | mean | 1.07 | 1.18 | 1.21 | 1.3 | 1.31 | 0.900 |
| Rate health excellent | % | 9.4 | 9.3 | 2.3 | 7.4 | 10.9 | 0.940 |
| Dementia before pandemic | % | 6.3 | 16.7 | 18.2 | 29.6 | 26.1 | 0.960 |
| Eat oily fish >=2x/wk | % | 7.8 | 18.5 | 22.7 | 29.6 | 30.4 | 1.000 |
| Smoker | % | 20.3 | 11.1 | 20.5 | 7.4 | 8.7 | 1.000 |
| College degree | % | 15.6 | 27.8 | 35.4 | 37 | 37 | 1.000 |
| Blood pressure medications | % | 4.7 | 7.4 | 9.1 | 3.7 | 6.5 | 1.000 |
| MI before baseline | % | 6.3 | 7.4 | 9.1 | 11.1 | 13 | 1.000 |
| MI before pandemic | % | 12.5 | 18.5 | 15.9 | 14.8 | 15.2 | 1.000 |
| Stroke before baseline | % | 4.7 | 3.7 | 2.3 | 0 | 2.2 | 1.000 |

**Supplemental Table 6. Data sources**

| Variable | Source documentation |
|---|---|
| Covariates | |
| Sex | https://biobank.ndph.ox.ac.uk/showcase/field.cgi?id=31 |
| Race | https://biobank.ndph.ox.ac.uk/showcase/field.cgi?id=21000 |
| Age (on January 1, 2020)[1] | https://biobank.ndph.ox.ac.uk/showcase/field.cgi?id=34 |
| Years since enrollment[2] | https://biobank.ndph.ox.ac.uk/showcase/field.cgi?id=53 |
| Estimated Omega-3 Index (%) | Schuchardt et al. (2022) |
| Plasma DHA (% of total fatty acids) | https://biobank.ndph.ox.ac.uk/showcase/field.cgi?id=23457 |
| Waist Circumference | https://biobank.ndph.ox.ac.uk/showcase/field.cgi?id=48 |
| BMI (kg/m$^2$) | https://biobank.ndph.ox.ac.uk/showcase/field.cgi?id=21001 |
| Townsend Deprivation Index | https://biobank.ndph.ox.ac.uk/showcase/field.cgi?id=189 |
| Education - College/University | https://biobank.ndph.ox.ac.uk/showcase/field.cgi?id=6138 |
| Smoking status - Never | https://biobank.ndph.ox.ac.uk/showcase/field.cgi?id=1249 |
| Self-reported health - Excellent | https://biobank.ndph.ox.ac.uk/showcase/field.cgi?id=2178 |
| Long standing illness- None | https://biobank.ndph.ox.ac.uk/showcase/field.cgi?id=2188 |
| Cystatin C | https://biobank.ndph.ox.ac.uk/showcase/field.cgi?id=30720 |
| Vitamin D | https://biobank.ndph.ox.ac.uk/showcase/field.cgi?id=30890 |
| HBA1c | https://biobank.ndph.ox.ac.uk/showcase/field.cgi?id=30750 |
| FEV | https://biobank.ndph.ox.ac.uk/showcase/field.cgi?id=3063 |
| Omega6 | https://biobank.ndph.ox.ac.uk/showcase/field.cgi?id=23452 |
| BP meds | https://biobank.ndph.ox.ac.uk/showcase/field.cgi?id=6177 |
| HDL-C | https://biobank.ndph.ox.ac.uk/showcase/field.cgi?id=23406 |
| Diabetes – yes | https://biobank.ndph.ox.ac.uk/showcase/field.cgi?id=2443 |
| Walking pace - Slow | https://biobank.ndph.ox.ac.uk/showcase/field.cgi?id=924 |
| Fresh fruit | https://biobank.ndph.ox.ac.uk/showcase/field.cgi?id=1309 |
| Dried fruit | https://biobank.ndph.ox.ac.uk/showcase/field.cgi?id=1319 |
| Salad/fresh vegetables | https://biobank.ndph.ox.ac.uk/showcase/field.cgi?id=1299 |
| Cooked vegetables | https://biobank.ndph.ox.ac.uk/showcase/field.cgi?id=1289 |
| Grain fiber | https://biobank.ndph.ox.ac.uk/showcase/field.cgi?id=1438<br>https://biobank.ndph.ox.ac.uk/showcase/field.cgi?id=1448<br>https://biobank.ndph.ox.ac.uk/showcase/field.cgi?id=1458<br>https://biobank.ndph.ox.ac.uk/showcase/field.cgi?id=1468<br>Using the approach here<br>https://www.ncbi.nlm.nih.gov/pmc/articles/PMC5799609/<br>to compute grain fiber grams/week |
| Outcomes | |
| Positive test[3] | https://www.ukbiobank.ac.uk/enable-your-research/about-our-data/covid-19-data |
| Hosp due to COVID[3] | https://www.ukbiobank.ac.uk/enable-your-research/about-our-data/covid-19-data |
| Dead from COVID[3] | https://www.ukbiobank.ac.uk/enable-your-research/about-our-data/covid-19-data |

1. Computed as the difference between month/year of birth and Jan 2020.

2. Computed as the difference between enrollment date and Jan 1, 2020.
3. Data for this project was available via the emergency use COVID-19 research data portal available to UKBiobank researchers through the end of 2021. Continuing data is available via standard UKBiobank data access portals.
